# Large-scale, in-house production of viral transport media to support SARS-CoV-2 PCR testing in a multi-hospital healthcare network during the COVID-19 pandemic

**DOI:** 10.1101/2020.04.29.20085514

**Authors:** Kenneth P. Smith, Annie Cheng, Amber Chopelas, Sarah DuBois-Coyne, Ikram Mezghani, Shade Rodriguez, Mustafa Talay, James E. Kirby

**Author notes:** These authors contributed equally to this work (Amber Chopelas, Sarah DuBois-Coyne, Ikram Mezghani, Shade Rodriguez, Mustafa Talay). Corresponding Author James E. Kirby, Beth Israel Deaconess Medical Center, 330 Brookline Avenue - YA309, Boston, MA 02215.

## Abstract

The COVID-19 pandemic has severely disrupted worldwide supplies of viral transport media (VTM) due to widespread demand for SARS-CoV-2 RT-PCR testing. In response to this ongoing shortage, we began production of VTM in-house in support of diagnostic testing in our hospital network. As our diagnostic laboratory was not equipped for reagent production, we took advantage of space and personnel that became available due to closure of the research division of our medical center. We utilized a formulation of VTM described by the CDC that was simple to produce, did not require filtration for sterilization, and used reagents that were available from commercial suppliers. Performance of VTM was evaluated by several quality assurance measures. Based on Ct values of spiking experiments, we found that our VTM supported highly consistent amplification of the SARS-CoV-2 target (coefficient of variation = 2.95%) using the Abbott RealTime SARS-CoV-2 EUA assay on the Abbott m2000 platform. VTM was also found to be compatible with multiple swab types and, based on accelerated stability studies, able to maintain functionality for at least four months at room temperature. We further discuss how we met logistical challenges associated with large-scale VTM production in a crisis setting including use of staged, assembly line for VTM transport tube production.

## Introduction

The COVID-19 pandemic has led to an unprecedented need for diagnostic RT-PCR testing. The ideal specimen type is currently believed to be a nasopharyngeal (NP) swab specimen transported to a molecular microbiology laboratory in viral transport medium (VTM). Starting in March 2020, increasing demand for testing led to a national shortage of both NP swabs and VTM that created significant bottlenecks in large-scale testing efforts. We discuss a nationwide collaborative effort produce 3-D printed swabs to address the former bottleneck in a separate manuscript (1). Here, we describe processes for large-scale, local production of VTM in large scale within the framework of a rigorous quality assurance program as a model to address unmet need for diagnostic supplies in a crisis setting.

VTM exists in several formulations, all of which consist of a buffered salt solution, a complex source of protein and/or amino acids, and antimicrobial agents. Its purpose is to preserve virus for later amplification by NAAT technology and/or viral culture. Although simpler formulations, for example, saline, are technically compatible with RT-PCR, most NAAT assays for respiratory pathogens have been developed and FDA-cleared for use with more complex transport media (i.e. VTM and universal transport medium, UTM). Further, stability of virus in saline over time may not be ideal, leading to degradation of viral nucleic acid, a particular concern for single-stranded RNA viruses such as SARS-CoV-2, and ultimately compromise detection. Overgrowth of bacteria may also occur in media lacking antimicrobial agents. As such, we chose to reproduce a standard of care transport medium to serve our healthcare network.

## Materials and Methods

### VTM Preparation

VTM was prepared using CDC standard operating procedure DSR-052–01 (2) with Hank’s balanced salt solution (HBSS) including phenol red (Gibco, Waltham, MA or Millipore, Burlington, MA) serving as this medium’s balanced salt solution which was supplemented with fetal bovine serum (FBS, Gibco or Corning, Tewksbury, MA), amphotericin B (Hyclone, Waltham MA, Corning, or Sigma, Burlington, MA), and gentamicin (Hyclone, Corning, or Sigma) to concentrations of 2%, 0.5 μg ml^−1^, and 100 μg ml^−1^, respectively. Component solutions were sterile and all liquid handling steps were performed using sterile technique in a class II biosafety cabinet to ensure sterility of the final VTM. Biosafety cabinets were thoroughly wiped down with 70% ethanol and UV decontaminated before and after use.

A foot pedal or hand controlled peristaltic pump was used to dispense media (Flexipump, Interscience, Woburn, MA or Digital MiniPump, Argos Technologies, Vernon Hills, IL). Sterilization of pump tubing was achieved by continuous dispensing of ~150 ml of a 10% bleach solution. Tubing was cleared of bleach by two extensive wash steps, each using a different bottle of sterile distilled water. Immediately before aliquoting, the pump was primed by dispensing at least 150 ml of VTM. After sterilization and priming, 3 ml of media was aliquoted into conical 15 ml centrifuge tubes (Falcon or Corning).

### Quality control (QC)

Each day’s production of VTM was assigned a new lot number for independent assessment in our quality control program to address any potential variability introduced by sterilization of our semi-automated pumping apparatus. Randomly selected tubes (at least 5 per lot) were examined to confirm appropriate media volume, color, optical clarity, and integrity of the tubes and caps. Next, one ml of media from a randomly selected tube in each lot was also plated on chocolate agar. The chocolate agar plate was dried in a biosafety cabinet and incubated overnight at 35°C to evaluate VTM sterility.

Finally, a randomly selected tube of media from each lot was tested for its support of SARS-CoV-2 RT-PCR testing using the EUA-authorized Abbott RealTime SARS-CoV-2 assay run on the Abbott m2000 platform, the same system used for clinical testing in our healthcare network. Accuplex COVID-19 reference material (SeraCare, Milford, MA), consisting of recombinant Sindbis virus containing SARS-CoV-2 RNA amplicon target, was spiked into samples of VTM from the lot under study at 2x the limit of detection of the assay (200 copies/mL) to assess amplification of SARS-CoV-2 near the assay limit of detection. QC was considered to pass if both the internal control and SARS-CoV-2 were amplified with cycle threshold (Ct) values within acceptable limits reflecting the inherent small native variation of this assay. All spiked VTM Ct values were recorded and visualized in aggregate as a Levy-Jennings plot using GraphPad Prism 7.0. Potential contamination of VTM with SARS-CoV-2 amplicon was also evaluated by testing the VTM lot alone. This QC assessment was considered to have passed if the assay internal control (IC) demonstrated appropriate levels of amplification in the absence of SARS-CoV-2 detection

### Alternative swab and media testing

VTM was evaluated for compatibility with various swab types including NP swabs as well as non-standard swab types that we considered might be used in place of NP swabs to respond to shortages. Briefly, swabs were removed from packaging, broken or cut with office scissors if no breakpoint was present, and placed in VTM. This step was performed on an open bench with non-sterile gloved hands to approximate the clinical environment. Swabs in VTM were incubated for 16–24 hours at 4°C to mimic conditions during specimen transport, after which RT-PCR QC was performed as outlined in the QC section. Alternative media and swab combinations were tested in an identical manner. See Table 1 for a listing of alternative swabs tested in VTM and Table 2 for a listing of alternative swab/media combinations.

**Table 1.** Swab validation using in-house VTM.

| Swab type | Manufacturer | Positive Control | Negative Control |
| --- | --- | --- | --- |
| E-Swab | Beckton Dickinson | 25.07 | Not Detected |
| Foam applicator | Puritan | 25.11 | Not Detected |
| Female cleaning swab | Hologic | 25.37 | Not Detected |
| Urethral swab | Hologic | 24.77 | Not Detected |
| FLOQSwab | Copan | 25.63 | Not Detected |
| NP Swab | Diagnostic Hybrids | 24.38 | Not Detected |
| Disposable sampling swab | Miraclean Technologies | 25.50 | Not Detected |
| Lesion/Other Swab | Diagnostic Hybrids | 25.68 | Not Detected |

**Table 2.** Alternative media and swab combinations tested.

| Media | Swab | Manufacturer | Positive Control | Negative Control |
| --- | --- | --- | --- | --- |
| 0.9% saline | None | TekNova | 24.54 | Not Detected |
| 0.9% saline | None | Becton Dickinson | 24.59 | Not Detected |
| 0.9% phosphate buffered saline (PBS) | None | Corning | 25.02 | Not Detected |
| Aptima Transport Media | Female cleaning swab | Hologic | 24.87 | Not Detected |
| Liquid Amies | E-Swab | Becton Dickinson | 25.91 | Not Detected |
| Viral Collection Media | NP swab | Diagnostic Hybrids | 24.65 | Not Detected |

### Accelerated Stability Testing

Two randomly selected tubes of VTM were incubated at either 56°C or 4°C (CDC recommended storage temperature) for 12 days. After incubation, visual inspection was performed, and the suitability of aged VTM for RT-PCR testing was evaluated as outlined in the QC section.

Stability of antimicrobial agents was confirmed by a killing study using *Escherichia coli* ATCC 25922 and *Candida albicans* ATCC 90028. Organisms were grown overnight on blood agar *(Escherichia coli)* or Sabouraud dextrose agar *(Candida albicans)* and suspended in 0.85% NaCl to a density of 0.5 McFarland using a Vitek DensiChek handheld colorimeter. For each organism, the 0.5 McFarland suspension was diluted 1:10 into 0.85% NaCl, and 10 μl of this dilution was added to 250 μl of VTM. An aliquot was immediately plated onto media using the drop plate method to quantify the initial inoculum prior to significant antibiotic exposure (3).

Organisms suspended in VTM were then incubated for 24 hours at 4°C as might occur during normal specimen transport and storage in the laboratory prior to testing. Colony forming units were then quantified using the drop plate method, and the percent recovery after VTM incubation was determined. The metric part for acceptability for this part of the accelerated stability study was >99% killing of *E. coli* and *C. albicans* in VTM aged at elevated temperature.

## Results

### VTM formulation

In response to the COVID-19 pandemic, our molecular microbiology laboratory was able to quickly scale up SARS-CoV-2 RT-PCR testing to approximately 1000 tests per day. However, national shortages of collection materials including VTM were projected to limit our ability to continue testing at this level. Therefore, we implemented in-house production of VTM to support testing needs in our hospital network.

Broadly speaking, we had two potential options for VTM production: following a recipe released by the CDC or attempting to “reverse-engineer” a recipe to mimic commercial formulations. We selected the CDC VTM medium for several reasons. First, exact formulations of commercial proprietary VTM are difficult to obtain. In contrast, components of the CDC VTM are explicitly defined. Second, all reagents necessary for the CDC formulation are commonly used for cell culture and were already available in our research division and available to order. Third, these component reagents could be purchased as sterile products, eliminating the time consuming process and technical difficulties associated with sterile filtration of large quantities of media (>40 L per week), especially in light of concurrent shortage of filtration devices.

We modified the CDC recipe slightly by inclusion of 10 mg L^−1^ of phenol red. Phenol red is a pH indicator that is pink or red at neutral or basic pH and transitions to yellow at acidic pH, providing a visual check on media pH. Furthermore, phenol red turns purple in the presence of bleach, thereby allowing us to visually confirm that that our dispense tubing was appropriately washed after bleach sterilization.

### Infrastructure, team organization, and management

In-house production of VTM is a technical and logistical challenge requiring dedicated space and personnel. Our diagnostic laboratory’s staffing and infrastructure was fully committed to addressing diagnostic testing needs and could not accommodate the additional burden of VTM production. However, early in the pandemic, our institution discontinued all non-essential research activities, resulting in availability of space and personnel, which and who could be redeployed for this effort.

Therefore, two tissue culture rooms with 8 class II biosafety cabinets were repurposed for VTM production efforts. In addition, a team of 9 research personnel were redeployed to the VTM production team, all of whom had significant laboratory experience including sterile technique. These personnel were divided into independently functioning teams working in rotations, so that if any team became infected with SARS-CoV-2 the other teams would be able to continue production. Direct oversight of team personnel, supply chain management, and maintenance of the quality control/quality assurance program was delegated to a senior CPEP medical microbiology fellow under supervision of the clinical microbiology laboratory director.

For VTM transport tube production, we established a staged assembly line taking advantage of the multiple available biosafety cabinets (Figure 1). Each cabinet was used sequentially for multiple production steps. The first biosafety cabinet was used as a staging area where conical tubes were uncapped and arranged in racks. Caps were stored for later use in sterile bags that originally held the tubes. The worker responsible for uncapping then shifted to the next empty biosafety cabinet to start the process again while a peristaltic pump was brought into proximity on a laboratory cart to fill uncapped tubes. After the tubes in the first biosafety cabinet were filled, a third worker capped the tubes, and finally a fourth worker bagged and boxed tubes for later transport to a packaging facility. This cycle proceeded continuously during production, with personnel moving sequentially to each of four biosafety cabinets to keep each step of the production line running. In this way, we were able to prepare 3500–4000 VTM tubes per day.

**Figure 1.**
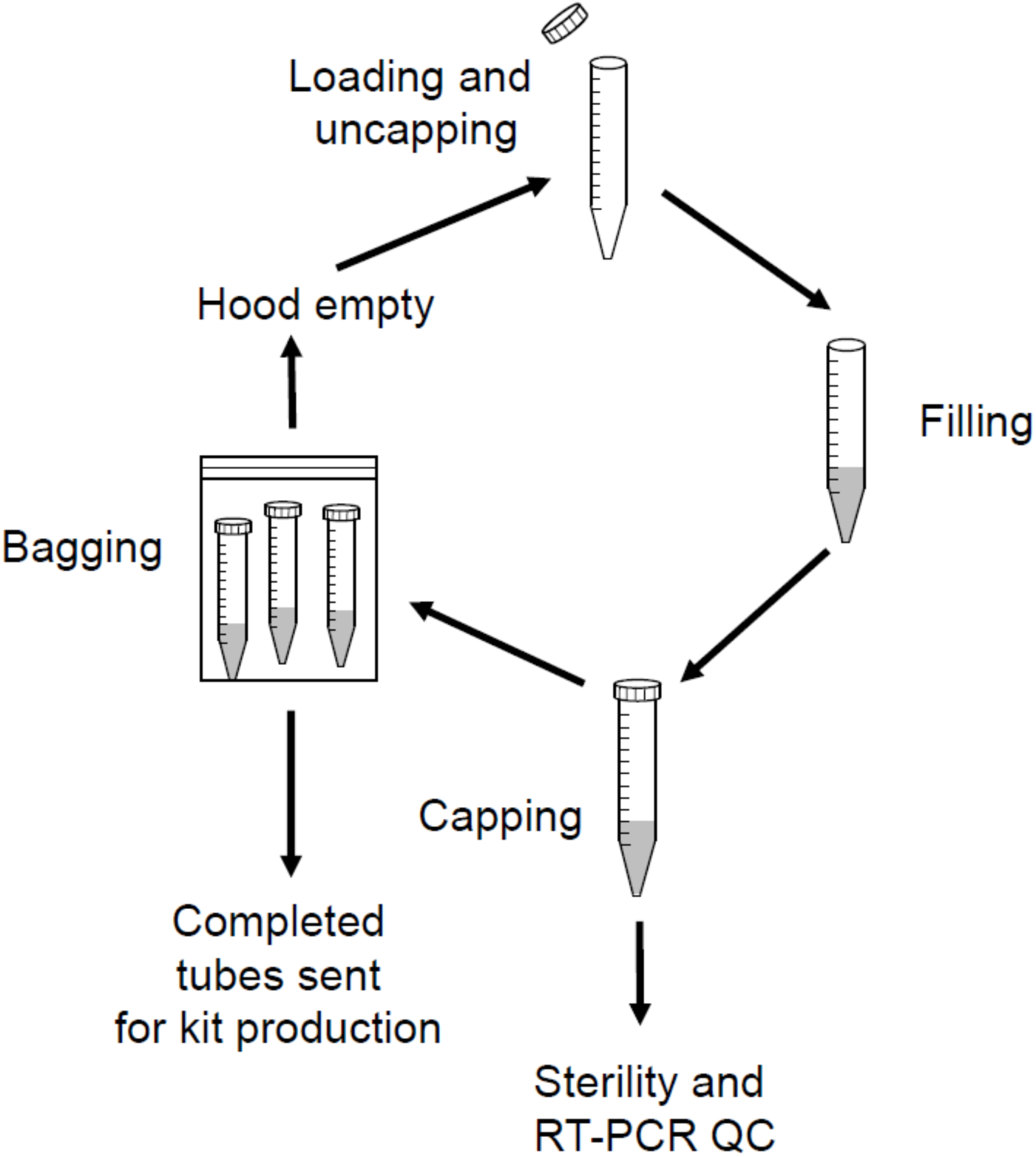
Workflow diagram. Each biosafety cabinet was used for all steps in the VTM production workflow. Personnel and the peristaltic pump rotated between cabinets but tubes remained in place until packaging. First, tubes were loaded and uncapped. Media filling was then accomplished through use of a peristaltic pump system moved to each biosafety cabinet in turn on a mobile cart. Filled tubes were capped and random samples were subject to QC. Tubes were then removed from the hood, bagged, and sent for distribution. The now empty hood was then used to start the next production cycle.

**Figure 2.**
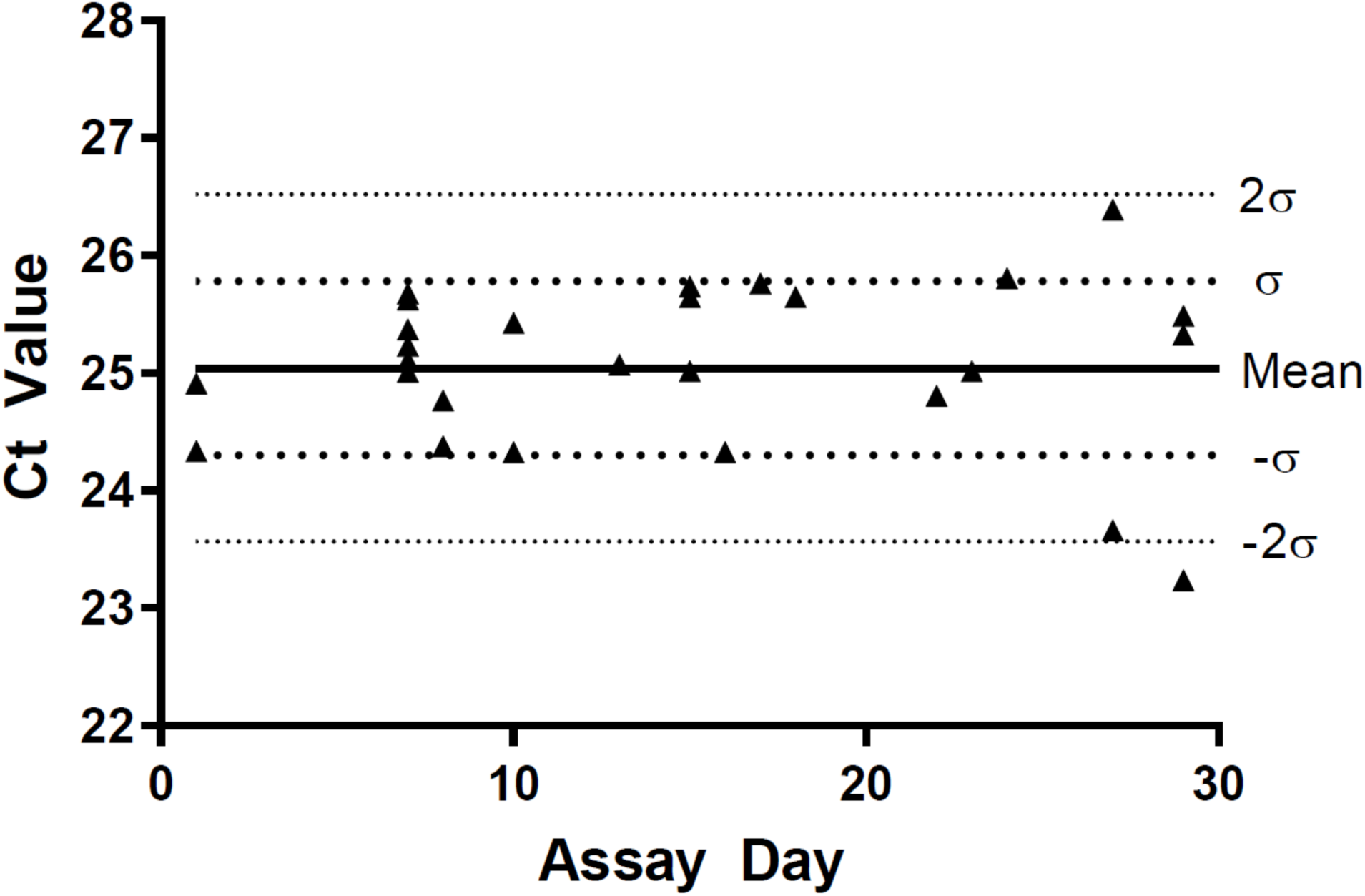
Levy-Jennings plot of VTM quality control data. Ct values for VTM lots spiked at 2X LoD with SARS-CoV-2 target were plotted each day of testing. Test dates with more than one data point represent the same batch of VTM evaluated for compatibility with multiple swab types (see Table 1).

Importantly, it should also be noted that releasing VTM for clinical use requires collaboration with personnel outside the laboratory. A separate team of 8–12 personnel in our materials management and distribution department were tasked with labeling each VTM tube, including with designated lot numbers, and pairing it with a NP collection swab to create a complete collection kit. This was an additional labor-intensive effort of critical importance.

### Quality control

Quality control was performed on each batch of VTM prior to release for clinical use. QC was incorporated into normal clinical workflow and was performed identically to patient specimens. Controls were spiked at 2x the limit of detection (LoD) of the assay, stated in the Abbott EUA documentation (200 copies mL^−1^) and independently confirmed in our internal verification studies, to enable detection of impactful deviations in the analytical sensitivity of the assay. In 27 quality control experiments thus far, representing > 50,000 VTM tubes, as well as swab compatibility studies, spiked SARS-CoV-2 amplicon was detected in all samples, indicating that the upper bound of deviation in LoD using home-made VTM was no more than 2-fold above the specified LoD and therefore PCR efficiency was no more than minimally affected if at all.

Furthermore, on average, observed Ct values were 25.0±0.7 for our spiked controls, approximately, one cycle below the mean Ct of 26.0±1.0 found for the LoD during our internal verification studies (n=80), and therefore, at the expected Ct value for a spiked sample at 2x LoD. The mean internal control Ct was 17.1±0.4 for the same reactions. We used coefficient of variation (CV) to evaluate relative variability of spiked target and internal control and found they were similar, at 3.0% and 2.4%, respectively. The CV for our LoD study was similar at 4.0% with slightly increased variance noted closer to the LoD as expected. We additionally visualized variance in spiked controls using as a Levy-Jennings plot (Figure 1). The majority of Ct values (96.2%) fell within ±2 standard deviations from the mean. A single value (3.8%) was >2 standard deviations from the mean, as is expected for 27 normally distributed data points. We did not observe any obvious trending in the data.

### Alternative swab and media testing

In light of the national shortage of NP swabs, it may become necessary to pair alternative swab types with in-house prepared VTM. Therefore, multiple swab types were qualitatively evaluated as previously described (1) and then incubated in our VTM at 4°C overnight. The VTM was then tested with the Abbott SARS-CoV-2 assay. Importantly, we found that all Ct values (Table 1) spiked with SARS-CoV-2 RNA at 2x the LoD fell within ±2 standard deviations of the mean previously determined in our QC studies of VTM without swabs, indicating compatibility of multiple swabs types with our prepared VTM.

### Accelerated stability testing

Unprecedented test volumes for SARS-CoV-2 resulted in a similarly unprecedented need for storage of collection kits. This is complicated by the CDC recommendation that VTM should be refrigerated. Clinical sites were not designed with large-volume refrigeration space. A priori, however, there was no obvious reason why VTM would require low temperature storage. Therefore, we sought to test whether the VTM formulation would remain stable at room temperature for prolonged periods.

In an emergent situation, real-time stability testing is impractical. Therefore, we performed an accelerated aging study using elevated temperature incubation (56°C) to predict effects of extended storage at room temperature (22°C). This was based on precedent in drug and diagnostic reagent stability testing for application of the Arrhenius equation (4, 5), which can be used to predict the effect of temperature on decay rate of reagents. Importantly, we found that PCR efficiency was maintained in accelerated-aged VTM based on internal control amplification, i.e., Ct of internal control remained within the expected mean and standard deviations of non-aged VTM. Antimicrobial ability to inhibit bacterial and fungal growth (gentamicin and amphotericin B) was also preserved. Based on Arrhenius equation calculations (4), maintenance of critical metrics during the tested 2 week incubation at 56°C was used to predict VTM stability for at least 4 months at room temperature.

## Discussion

Production of VTM on a scale capable of supporting a hospital network requires extensive infrastructure including bench space, reagent storage, and multiple biosafety cabinets, which are unavailable in routine clinical microbiology laboratories. Further, dedicated personnel with laboratory experience are required at a time when technologists are needed to address a surge in routine diagnostic work. Here, we took advantage of our medical center’s extensive research division, which was largely shuttered as a response to the pandemic, to repurpose resources for large-scale VTM tube production.

We acknowledge that saline has been advocated by the CDC as an alternative transport medium (6). However, none of our amplification methods were qualified in their EUA or FDA-clearance to be used with saline. Furthermore, there were theoretical advantages in terms of potentially sustaining viral integrity and presumably labile RNA viral genome target material through use of a buffered medium with a protein component for stabilization.

Another alternative that we considered was the use of a guanidinium-based transport buffer. It has advantage in stabilization of nucleic acid and inactivation of virus (7). We determined that the Abbott M2000 EUA was compatible with Aptima (Hologic Marlborough, MA) guanidinium-based transport buffer, and the guanidinium-based Abbott multi-Collect Specimen transport buffer (8) which is listed as an acceptable transport medium in the Abbott RealTime SARS-CoV-2 EUA instructions. However, guanidinium-based transport medium of any kind was not listed as options in the package inserts for either FDA-cleared respiratory PCR assays or SARS-CoV-2 EUA assays on ePlex (Genmark, Carlsbad, CA) and Cepheid (Danaher, Sunnyvale, CA) systems used in our hospital. As such, use of a non-standard viral transport media would have necessitated additional validations on both systems at a time when COVID-19 test reagents were on strict allocation. It was also uncertain whether such guanidinium-based medium would ultimately prove compatible with the specific chemistries of these platforms. Although not described here, we confirmed that our VTM was compatible with the GenMark ePlex respiratory viral panel, Cepheid influenza test, and SARS-CoV-2 tests on both platforms (data not shown).

Alternative VTM and recipes exist, but required weighing and solubilizing materials, followed by filter sterilization. A replication of commercial universal transport medium was tried but became untenable during scale-up. In contrast, all components for CDC VTM are available as sterile solutions, which streamlined the production process and helped ensure lot-to-lot consistency.

Although the CDC formulation of VTM requires only a few ingredients, initial production of VTM was a significant challenge as components were not stocked in the clinical laboratory and supply delivery during the crisis was unacceptably slow. Initially, therefore, donations of existing supplies were requested through broadcast email to our research community and beyond through social media until sufficient supplies could be obtained from outside vendors. The response was exceedingly positive, and supplies were provided in abundance with meticulous accounting for donations from research laboratories to later reimburse costs. Laboratory supply companies also quickly placed the reagent used to make VTM on allocation. Although this was meant to prioritize orders from hospitals over research laboratories, it also resulted in these reagents appearing out of stock when ordered through existing channels. We therefore needed to place each order by directly contacting company representatives to ensure stock would be appropriately released.

After solving logistical issues, we identified several procedural concerns, which initially seemed trivial but became significant hurdles at scale. One of these was tube filling. At capacity, our goal was to produce 3500 to 4000 tubes per day requiring handling of approximately 50 to 60L of media per week. Manual filling of this number of tubes is not practical. Therefore, we acquired and utilized a semi-automated peristaltic pump that repeatedly dispensed appropriate volumes. The pumps were controlled by a foot pedal or a push button on a dispensing gun, allowing repeatable delivery of 3 mL to each tube much more efficiently than manual pipetting.

Another example of a simple procedure that becomes complex at scale was tube capping. Each of our personnel routinely capped approximately 1,000 tubes per day. This is a demanding process, which invariably resulted in painful blisters and was therefore not sustainable. We also observed that Falcon 15 mL conical tubes, that were used initially, required substantial force to adequately close. Therefore, we switched to Corning tubes with Centristar caps which proved much easier to manipulate, but did not entirely solve the blistering issue. We therefore attempted to protect the fingers of personnel by wrapping them in different types of tape including surgical tape (Blenderm, 3M, Saint Paul, MN), self-adherent wrap (Coban, 3M), or KT blister prevention tape (KTTape, Linden, UT). Each proved suboptimal, either being too thin, too thick, or limiting dexterity. Ultimately, we identified rock climbing tape (a donation from Metolius Climbing, Bend, Oregon) as an ideal solution which eliminated blistering entirely so that capping was fully tolerated by staff.

Aside from logistical issues, we would like also to emphasize the quality control plan put in place. This included testing sterility and PCR efficiency for each lot through assessment of Ct variance of spiked SAR2-CoV-2 and the internal control, results which could be analyzed using in standard Levey-Jennings plots to detect bias and issues with reproducibility.

Taken together, our experience with VTM production provides an example of a rigorously controlled effort to provide a critical resource at scale in a crisis. Through collaboration between researchers, clinical microbiologists, and support/logistics personnel, we were able to circumvent the limitations of a routine diagnostic laboratory and address a significant bottleneck that would otherwise limit high volume COVID-19 testing capacity.

## Data Availability

All data are included in the manuscript.

## Acknowledgements

K.P.S. was supported by the National Institute of Allergy and Infectious Diseases of the National Institutes of Health under award number F32 AI124590. The content is solely the responsibility of the authors and does not necessarily represent the official views of the National Institutes of Health.

## Notes

### Competing Interest Statement

The authors have declared no competing interest.

